# Facility-based and virtual cardiac rehabilitation in young patients with heart disease during the COVID-19 era

**DOI:** 10.1101/2023.03.24.23287722

**Authors:** Elizabeth B. Aronoff, Clifford Chin, Alexander R. Opotowsky, Wayne A. Mays, Sandra K. Knecht, Jennah E. Goessling, Malloree C. Rice, Justine Shertzer, Samuel G. Wittekind, Adam W. Powell

## Abstract

**Background:** Cardiac rehabilitation (CR) is an important tool for improving fitness and quality of life in those with heart disease (HD). Few pediatric centers use CR to care for these patients, and virtual CR is rarely used. Additionally, it is unclear how the COVID-19 era has changed CR outcomes.

**Objectives:** This study assessed fitness improvements in young HD patients participating in both facility-based and virtual CR during the COVID-19 pandemic.

**Methods:** This retrospective single-center cohort study included new patients who completed CR from March 2020 through July 2022. CR outcomes included physical, performance, and psychosocial measures. Comparison between serial testing was performed with a paired t-test with P<0.05 was considered significant. Data are reported as mean±standard deviation.

**Results:** There were 47 patients (19±7.3 years-old; 49% male) who completed CR. Improvements were seen in peak oxygen consumption (VO_2_, 62.3±16.1 v 71±18.2% of predicted, p=0.0007), 6-minute walk (6MW) distance (401±163.8 v 480.7±119.2 meters, p=<0.0001), sit to stand (16.2±4.9 v 22.1±6.6 repetitions; p=<0.0001), Patient Health Questionnaire-9 (PHQ-9) (5.9±4.3 v 4.4±4.2; p=0.002), and Physical Component Score (39.9±10.1 v 44.9±8.8; p=0.002). Facility-based CR enrollees were less likely to complete CR than virtual patients (60%, 33/55 v 80%, 12/15; p=0.005). Increases in peak VO_2_ (60±15.3 v 70.2±17.8 % of predicted; p=0.002) were seen among those that completed facility-based CR; this was not observed in the virtual group. Both groups demonstrated improvement in 6MW distance, sit-to-stand repetitions, and sit-and-reach distance.

**Conclusions:** Completion of a CR program resulted in fitness improvements during the COVID-19 era regardless of location.

## Introduction

Cardiac rehabilitation (CR) has been established as a critical component in adult care but is rarely utilized in pediatric facilities. Children and adults with both congenital and acquired heart disease (HD) are often cared for in pediatric facilities during their entire lifespan^1^. Despite life-prolonging palliation for HD, children and adults with HD often have poor fitness that has been shown to decline over time on serial cardiopulmonary exercise testing (CPET)^2-5^. CR programs have started to emerge as a tool to reverse these negative fitness changes and to improve overall performance and quality of life ^6-9^. Despite this promising evidence, there remain few pediatric centers offering clinically based CR, thus there is limited “real world” evidence on the efficacy of clinical CR in young HD populations.

CR programs are traditionally facility-based, which can pose challenges related to frequent school and work absences and travel to facility-based sessions. There are a growing number of programs trying to develop virtual options for families, which has become even more important due to the COVID-19 pandemic and the need for disease mitigation strategies^10-11^. Virtual CR has been shown in small studies to be feasible and effective using home-based equipment^12^.

There has been minimal research on the utilization of virtual CR in young patients with HD. The aims of this study were to 1) assess physical and psychosocial outcomes of young HD patients participating in CR at our institution during the COVID-19 pandemic, and 2) compare these outcomes in those who completed facility-based versus virtual CR during that time.

## Methods

This is a retrospective cohort study assessing for change in patients undergoing either facility-based or virtual CR at our institution for the first time from March 2020 through July 2022. Data on enrollment dates, number of completed weeks and sessions, and information on program completion were collected. Additional data collected included patients’ demographic information and medical diagnosis. Exclusion criteria included previous enrollment in the CR program and failure to complete at least 12 weeks of the CR program. Patients who changed CR modality during the program (i.e., changing from facility-based to virtual) were included in the analysis of the entire cohort but excluded from the sub-analysis comparing facility-based to virtual CR.

CR is a personalized exercise program designed per the standard of the American Association of Cardiovascular and Pulmonary Rehabilitation (AACVPR)^13^. Participants were instructed to attend two to three sessions lasting 1 hour per week for 12 weeks. CR sessions either took place at our hospital’s cardiac rehabilitation gym, defined as facility-based or through a virtual encounter. Both programs included one-on-one supervision by a trained exercise physiologist. Each CR session consisted of 5–10 min of warm-up and stretching activity, 30 min of aerobic (facility-based) or high-intensity interval training (virtual), 15 min of low-resistance high-repetition strength training, and 5 min of cool-down activity. The aerobic training component was conducted to sustain 70–80% of the peak heart rate as assessed by baseline CPET. Age-appropriate activities were incorporated into sessions to attempt to increase motivation and enjoyment during CR. Participants were encouraged to remain active at home, but the non-CR-related physical activity level was not monitored, nor was an exercise prescription given during this phase of the CR program.

CR outcomes were recorded at the first and last CR sessions. These included physical body measurements, performance measures, and psychosocial questionnaires. Body composition was performed using the InBody bioimpedance analysis scale (InBody370; InBody, Cerritos, CA, USA) as previously described^14^. Waist, trunk, thigh, and mid-arm circumferences were measured. Cardiopulmonary exercise testing was performed on a stationary cycle ergometer (Corival; Lode; Groningen, The Netherlands) with an individualized ramp protocol, as previously described^14^. The rate of increase was chosen by experienced clinical exercise physiologists based on the patient’s body size and expected fitness level, targeting an exercise test duration of roughly 10 minutes. Cardiopulmonary responses to exercise were assessed using breath-by-breath analysis (Ultima CardiO2; MGC Diagnostics; Saint Paul, MN, USA). Criteria for a maximal effort exercise test were that 2 of the following 3 criteria were met: respiratory exchange ratio >1.10; maximal heart rate ≥85% of the age-predicted maximum (220-age in years); or maximal rating of perceived exertion >18 on a 6 to 20 Borg scale^15^. Predicted peak oxygen consumption (VO_2_ peak) was calculated per Wasserman et al. and Cooper et al. equations^16-17^.

Additional performance outcomes collected included sit-to-stand repetitions, sit-and-reach distance, arm curls, and a 6-minute walk test. Multiple social and emotional surveys were provided and completed pre and post-CR including the Patient Health Questionnaire-9 (PHQ-9) screen for depression, Duke Activity Status Index which assesses patient-reported functional status (higher score means higher functional status), and a Rate Your Plate score where patients rated the healthiness of their eating habits. Participants also completed the Mental Component Scoring (MCS) which represents the combination of the vitality, social functioning, role-emotional, and mental health forms, and the Physical Component Scoring (PCS), a combination of the physical function, role-physical, bodily pain, and general health forms. The MCS and PCS are components from the Short Form Health Inventory – 36^18-21^.

### Statistical Analyses

Descriptive statistics represented by means and standard deviations were calculated for each parameter. Baseline and CR completion data were compared using a paired T-test. Groups were compared at baseline and then at program completion utilizing Student’s t-test for independent groups with equal variances. All T-tests were two-sided with P value <0.05 considered significant. Statistical analyses were performed using JMP®, Version 16 from SAS Institute Inc. (Cary, NC). Figures were created with Microsoft Excel (Redmond, WA).

## Results

A total of 73 patients (55 facility-based, 15 virtual, 3 hybrids) enrolled in CR during the study period; 47 patients (64%) completed at least 12 weeks of the program. Patients enrolled in virtual CR had a higher completion rate than those enrolled in facility-based CR (80% (12/15 virtual participants) v 60% (33/55 facility-based participants, p=0.005 For facility-based CR, reasons for discontinuation were: 13 non-adherence without a specific reason, 7 underwent surgery, 1 intercurrent illness, and 1 medically unstable as rehabilitation progressed (pain and safety concerns related to an acute abdominal process). For virtual CR, the reasons for discontinuation were: 2 underwent surgery, and 1 due to family circumstances. The 2 patients who completed a hybrid facility-based/virtual CR (1 patient enrolled in hybrid but didn’t complete the program) were included in the total analysis but were excluded in the comparison between virtual and facility-based outcomes.

Demographics for the cohort are presented in Table 1. For CR, height (162.2±14.4 v 163.3±13.6 cm, p=0.005) and skeletal muscle mass (24.6±9.5 v 25.5±9.5 kg, p=0.002) increased (Table 2). Overall, there was a significant increase in peak VO_2_ (62.3±16.1 v 71±18.2 % of predicted, p=0.0008; 22.6±7.6 v 25.6±7.8 mL/kg/min, p=0.003), 6-minute walk distance (404.0±113.8 v 480.7±119.2 meters, p=<0.0001), sit to stand (16.2±4.9 v 22.1±6.6 repetitions; p=<0.0001), and arm curls (18.3±4.1 v 22.7±6.3 repetitions, p=<0.0001). Lastly, the cohort had a significant improvement in their PHQ-9 (5.9±4.3 v 4.4±4.2, p=0.02), Duke Activity Index scores (33.7±16.2 v 38.0±14.3, p=0.008), and PCS (39.9±10.1 v 44.9±8.8, p=0.002).

**Table 1:**
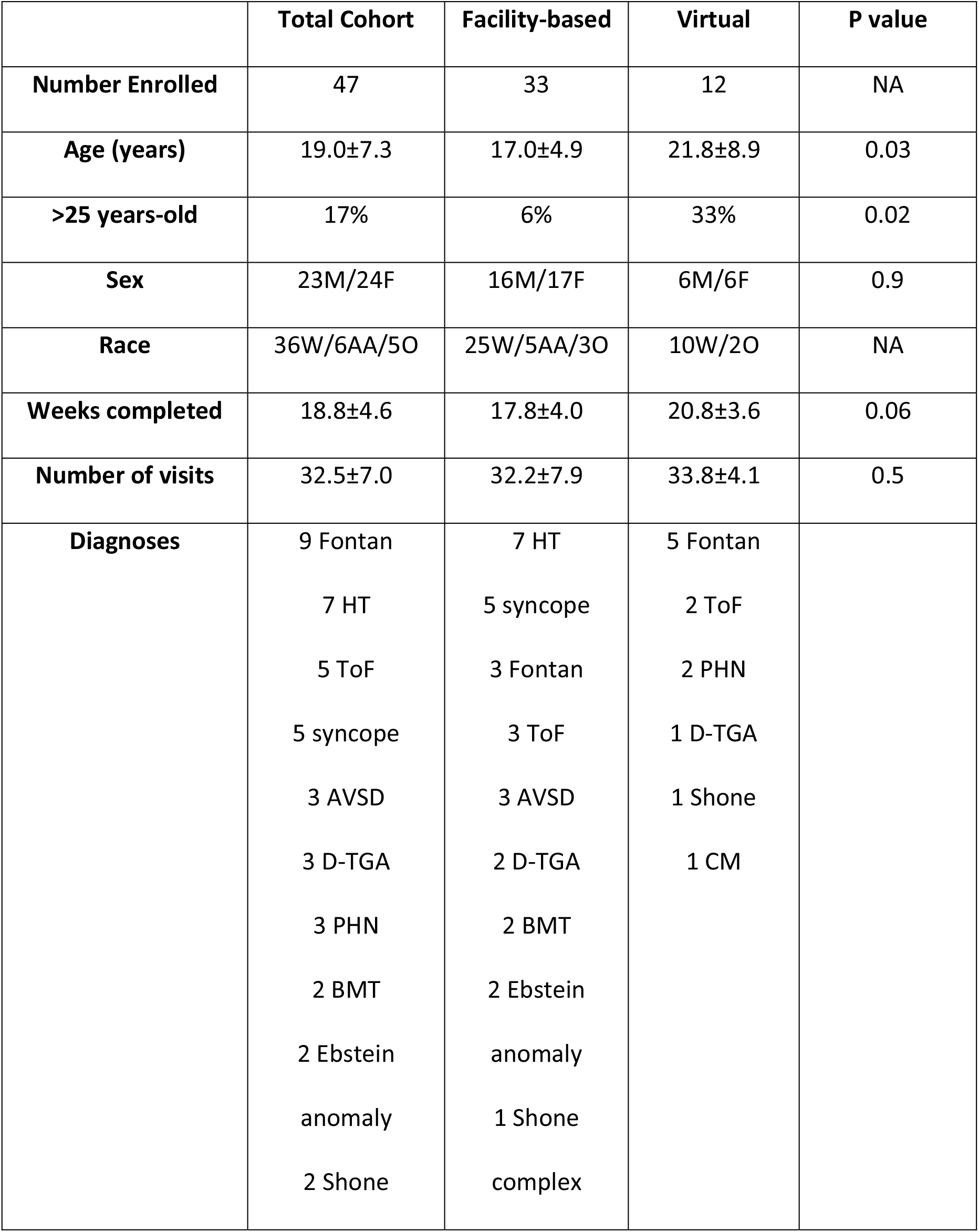
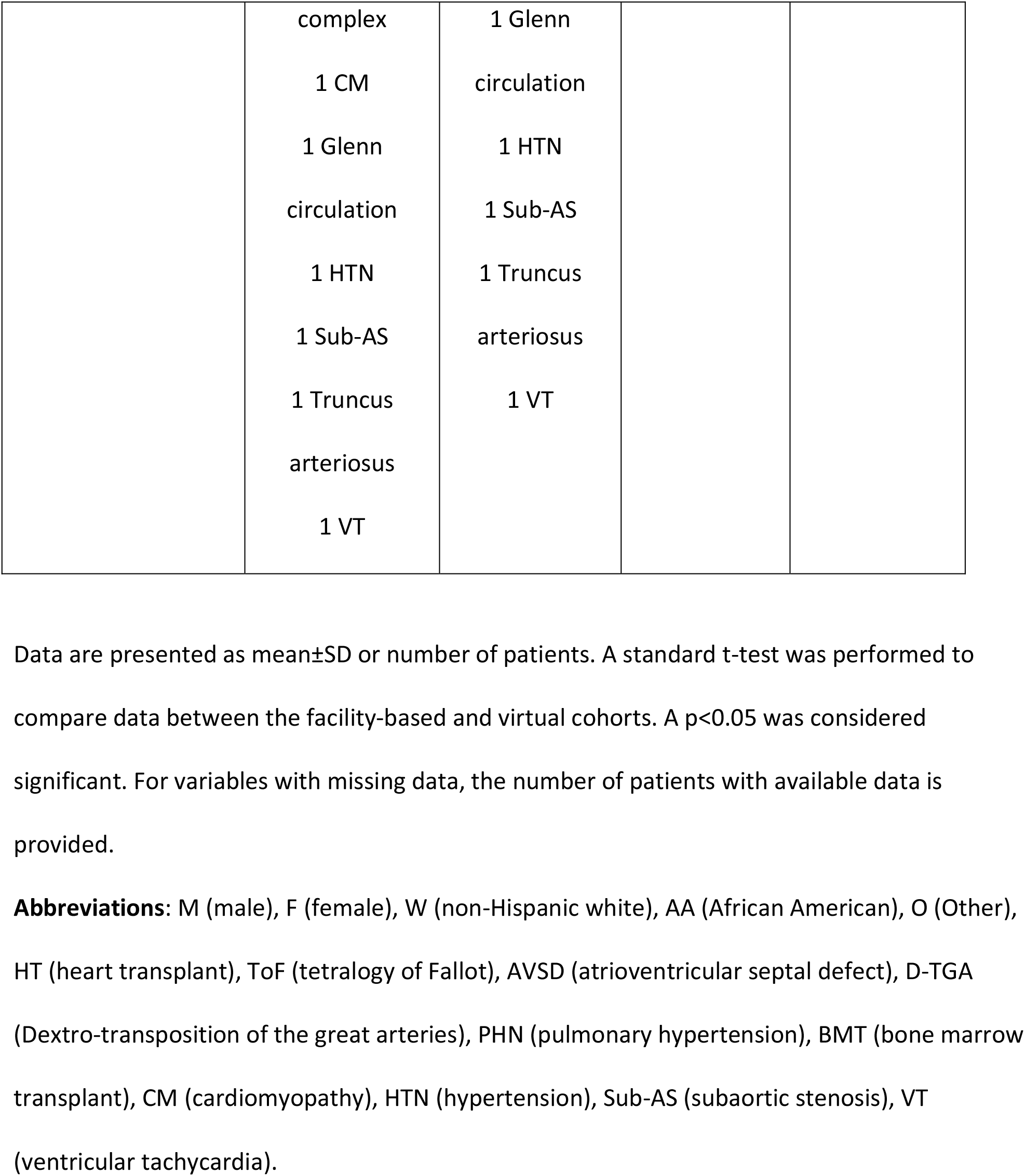
Baseline demographics of those who completed cardiac rehabilitation, including the total cohort and those that completed facility-based and virtual rehabilitation.

**Table 2:**
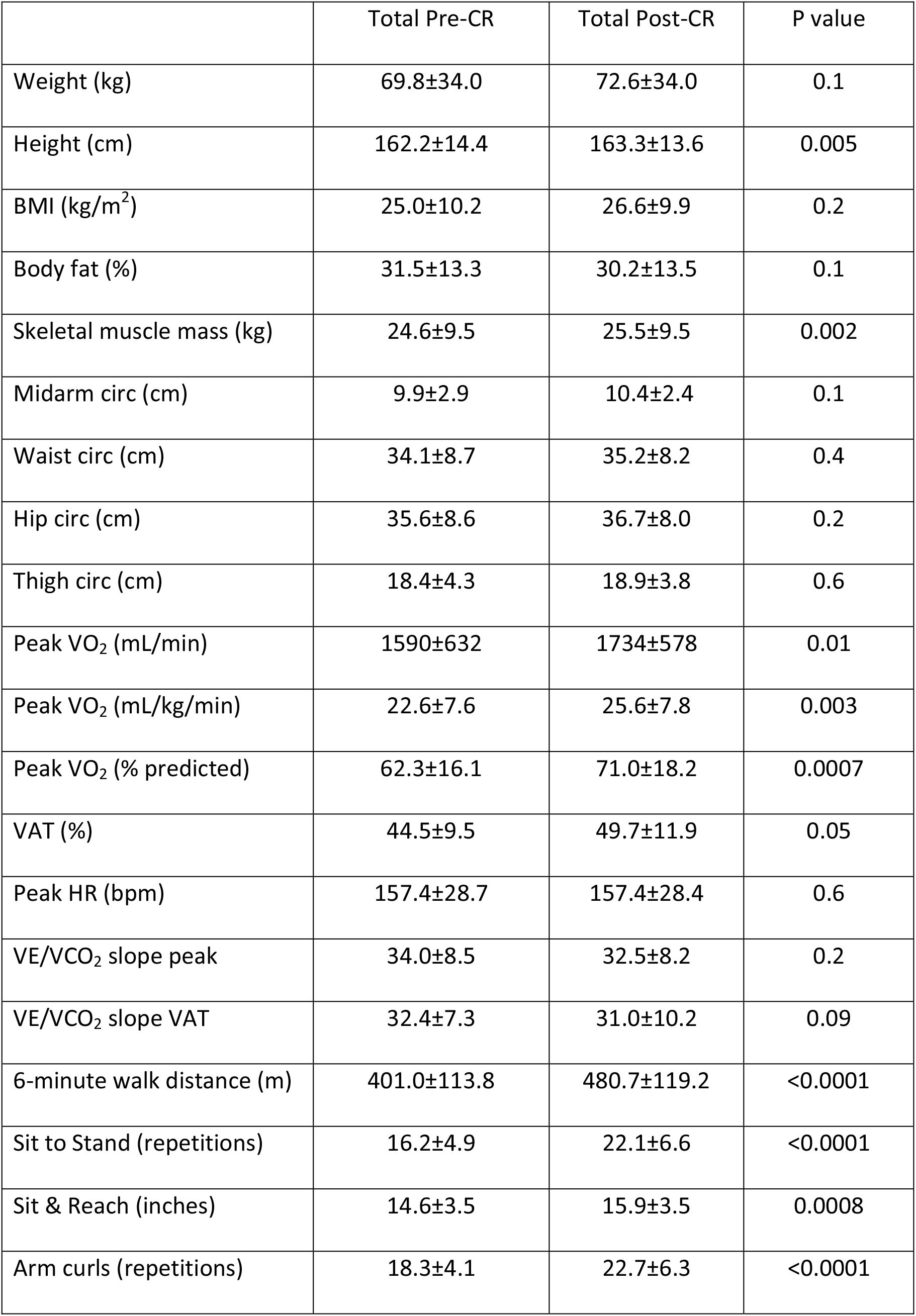
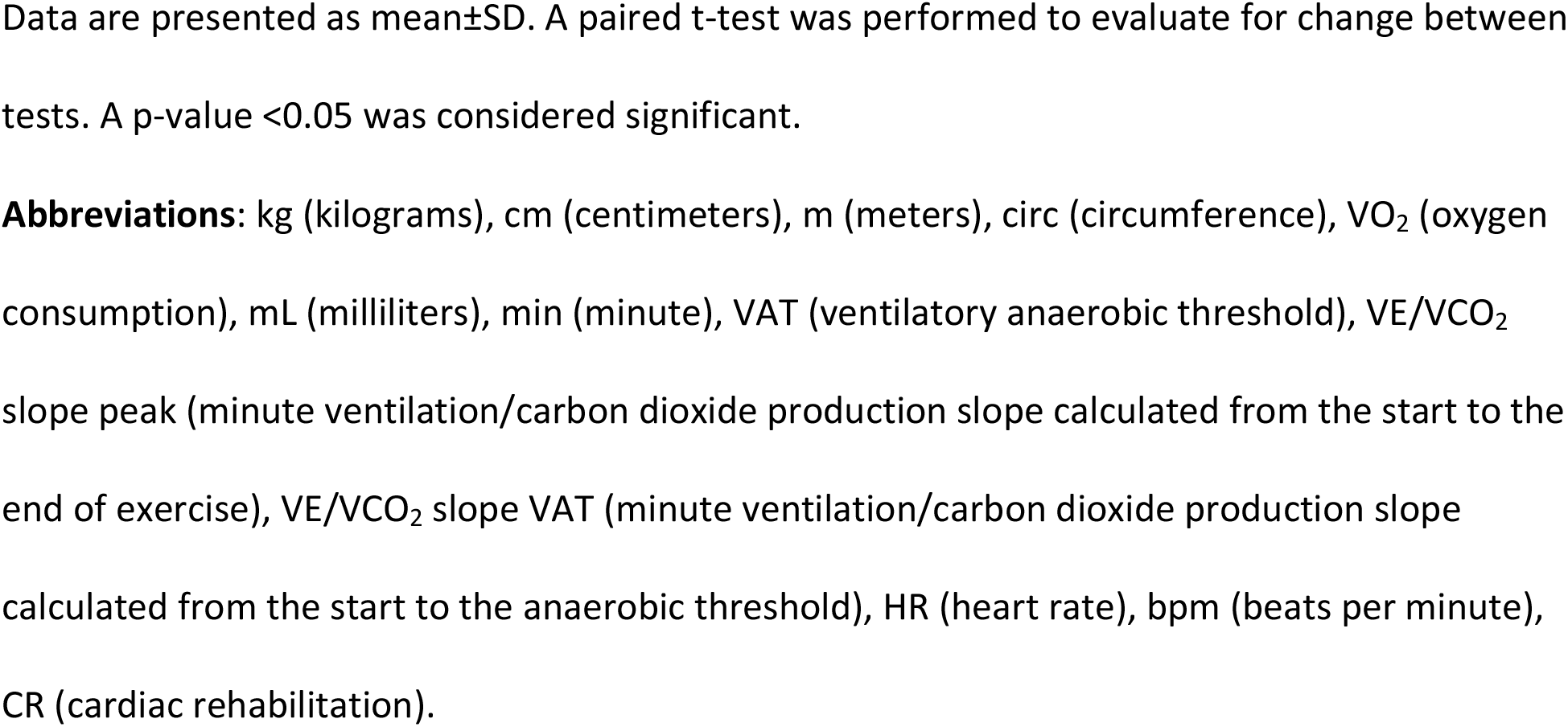
Body composition and fitness outcomes for the total cohort (n=47) pre- and post-cardiac rehabilitation.

|  | Total Pre-CR | Total Post-CR | P value |
| --- | --- | --- | --- |
| Weight (kg) | 69.8±34.0 | 72.6±34.0 | 0.1 |
| Height (cm) | 162.2±14.4 | 163.3±13.6 | 0.005 |
| BMI (kg/m<sup>2</sup>) | 25.0±10.2 | 26.6±9.9 | 0.2 |
| Body fat (%) | 31.5±13.3 | 30.2±13.5 | 0.1 |
| Skeletal muscle mass (kg) | 24.6±9.5 | 25.5±9.5 | 0.002 |
| Midarm circ (cm) | 9.9±2.9 | 10.4±2.4 | 0.1 |
| Waist circ (cm) | 34.1±8.7 | 35.2±8.2 | 0.4 |
| Hip circ (cm) | 35.6±8.6 | 36.7±8.0 | 0.2 |
| Thigh circ (cm) | 18.4±4.3 | 18.9±3.8 | 0.6 |
| Peak VO<sub>2</sub> (mL/min) | 1590±632 | 1734±578 | 0.01 |
| Peak VO<sub>2</sub> (mL/kg/min) | 22.6±7.6 | 25.6±7.8 | 0.003 |
| Peak VO<sub>2</sub> (% predicted) | 62.3±16.1 | 71.0±18.2 | 0.0007 |
| VAT (%) | 44.5±9.5 | 49.7±11.9 | 0.05 |
| Peak HR (bpm) | 157.4±28.7 | 157.4±28.4 | 0.6 |
| VE/VCO<sub>2</sub> slope peak | 34.0±8.5 | 32.5±8.2 | 0.2 |
| VE/VCO<sub>2</sub> slope VAT | 32.4±7.3 | 31.0±10.2 | 0.09 |
| 6-minute walk distance (m) | 401.0±113.8 | 480.7±119.2 | <0.0001 |
| Sit to Stand (repetitions) | 16.2±4.9 | 22.1±6.6 | <0.0001 |
| Sit & Reach (inches) | 14.6±3.5 | 15.9±3.5 | 0.0008 |
| Arm curls (repetitions) | 18.3±4.1 | 22.7±6.3 | <0.0001 |

Data are presented as mean±SD. A paired t-test was performed to evaluate for change between tests. A p-value <0.05 was considered significant.
**Abbreviations:** kg (kilograms), cm (centimeters), m (meters), circ (circumference), VO<sub>2</sub> (oxygen consumption), mL (milliliters), min (minute), VAT (ventilatory anaerobic threshold), VE/VCO<sub>2</sub> slope peak (minute ventilation/carbon dioxide production slope calculated from the start to the end of exercise), VE/VCO<sub>2</sub> slope VAT (minute ventilation/carbon dioxide production slope calculated from the start to the anaerobic threshold), HR (heart rate), bpm (beats per minute), CR (cardiac rehabilitation).

**Table 3:** Self-reported mental and social outcomes for the total cohort (n=47) pre- and post-cardiac rehabilitation.

|  | Total Pre-CR | Total Post-CR | P value |
| --- | --- | --- | --- |
| PHQ-9 | 5.9±4.3 | 4.4±4.2 | 0.04 |
| Duke Activity Score | 33.7±16.2 | 38.0±14.3 | 0.006 |
| MCS | 48.4±10.1 | 50.1±9.3 | 0.5 |
| PCS | 39.9±10.1 | 44.9±8.8 | 0.002 |
| Rate your plate | 45.5±8.6 | 47.3±8.1 | 0.06 |

Data are presented as mean±SD. A paired t-test was performed to evaluate for change between tests. A p-value <0.05 was considered significant.
**Abbreviations:** PHQ-9 (patient health questionnaire-9), MCS (mental component scoring for vitality, social functioning, emotion health, and mental health), PCS (physical component scoring for physical function, bodily pain, and general physical health).

On a further sub-analysis of the whole cohort, when excluding patients with either vasovagal syncope (n=5) or systemic hypertension (n=1), the remaining patients (n=41) showed an improvement in percent predicted peak VO_2_ (62.7±15.8 v 67.8±15.8%, p=0.02), 6-minute walk distance (392.2±113.5 v 484.3±111.9 meters, p<0.0001), sit to stand (16.2±4.9 v 21.1±5.9 repetitions, p<0.0001), sit and reach (14.2±3.6 vs 15.2±3.4 inches, p=0.01), PCS (40.5±9.9 v 44.7±9.3, p=0.03), and Duke activity score (31.9±16.4 v 36.4±14.5, p=0.03). There was no significant change in the PHQ-9 or MCS when focusing only on patients with intracardiac disease. For those under 18 years old (n=27), percent predicted peak VO_2_ increased (61.8±15.9 v 72.9±19.9%, p=0.002), as did 6-minute walk distance (382.8±133.3 v 463.8±137.5 meters, p<0.0001), sit to stand (17.7±5.3 v 22.9±7.1 repetitions, p=0.0003), sit and reach (15.0±3.9 v 16.3±4.0 inches, p=0.009), PCS (38.6±11.1 v 43.9±9.2, p=0.01), and Duke activity score (31.7±16.5 v 36.3±14.9, p=0.04). There was not a significant change in PHQ-9 and Rate your Plate.

When comparing the facility-based and virtual CR groups, the baseline and completion values between the facility-based and virtual cohorts were similar, although the virtual group tended to be older (facility-based 17.0±4.9 years v virtual 21.8±8.9 years, p=0.03) (Table 1; Supplemental Table 2). Throughout CR, the facility-based group had an increase in height (161.0±14.8 v 161.9±13.9 cm, p=0.04), body mass index (25.6±10.9 v 26.8±11.0 kg/m^2^, p=0.03), and skeletal muscle mass (23.6±9.9 v 24.5±10.1 kg, p=0.01) (Table 2). There was no significant difference in body composition on serial bioimpedance measurements in the virtual group. There was no significant change in body circumference measurements in either group. When evaluating the physical performance outcomes for the facility-based and virtual groups, the facility-based group demonstrated a statistically significant change in peak VO_2_ (60.0±15.3 v 70.2±17.8 % of predicted, p=0.01; 22.3±7.1 v 26.1±6.8 mL/kg/min, p=0.0008) without a change in the heart rate response (Table 4). There were no significant changes in any of the CPET parameters measured on serial testing in the virtual group. There were significant changes for both groups in the 6-minute walk (facility-based 384.5±118.7 v 488.2±12.5 meters, p=<0.0001; virtual 457.5±89.7 v 496.5±64.8 m, p=0.04), sit to stand (facility-based 15.8±5.0 v 22.1±6.9 repetitions, p=<0.0001; virtual 17.8±4.9 v 21.6±6.6 repetitions, p=0.03), sit and reach (facility-based 14.7±3.3 v 16.1±3.6 inches, p=0.004; virtual 14.4±4.2 v 15.0±3.3 cm, p=0.004), and arm curls (facility-based 17.9±4.4 v 23.0±5.1 repetitions, p=<0.0001; virtual 19.5±3.7 v 24.8±4.8 repetitions, p=0.005).

**Table 4:**
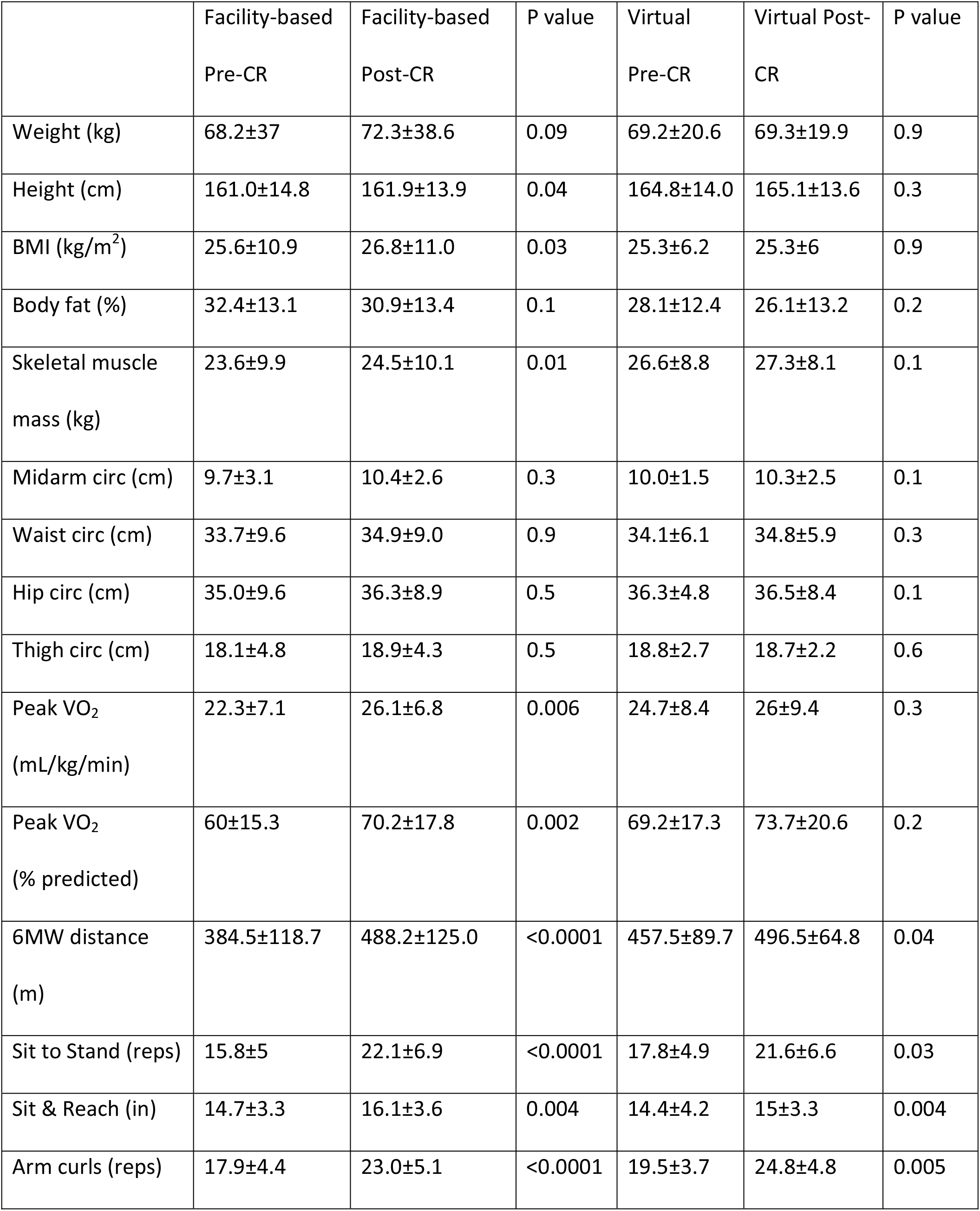
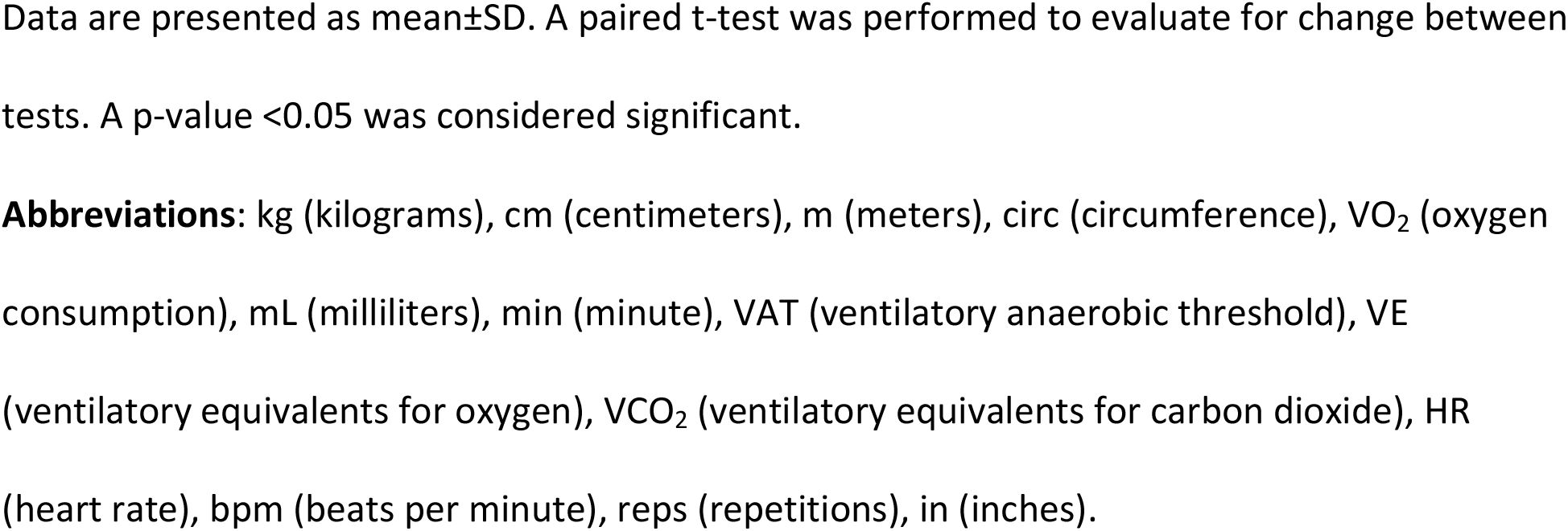
Body composition and fitness outcomes for the patients that completed facility-based (n=33) and virtual (n=12) cardiac rehabilitation.

|  | Facility-based<br>Pre-CR | Facility-based<br>Post-CR | P value | Virtual<br>Pre-CR | Virtual Post-CR | P value |
| --- | --- | --- | --- | --- | --- | --- |
| Weight (kg) | 68.2±37 | 72.3±38.6 | 0.09 | 69.2±20.6 | 69.3±19.9 | 0.9 |
| Height (cm) | 161.0±14.8 | 161.9±13.9 | 0.04 | 164.8±14.0 | 165.1±13.6 | 0.3 |
| BMI (kg/m<sup>2</sup>) | 25.6±10.9 | 26.8±11.0 | 0.03 | 25.3±6.2 | 25.3±6 | 0.9 |
| Body fat (%) | 32.4±13.1 | 30.9±13.4 | 0.1 | 28.1±12.4 | 26.1±13.2 | 0.2 |
| Skeletal muscle<br>mass (kg) | 23.6±9.9 | 24.5±10.1 | 0.01 | 26.6±8.8 | 27.3±8.1 | 0.1 |
| Midarm circ (cm) | 9.7±3.1 | 10.4±2.6 | 0.3 | 10.0±1.5 | 10.3±2.5 | 0.1 |
| Waist circ (cm) | 33.7±9.6 | 34.9±9.0 | 0.9 | 34.1±6.1 | 34.8±5.9 | 0.3 |
| Hip circ (cm) | 35.0±9.6 | 36.3±8.9 | 0.5 | 36.3±4.8 | 36.5±8.4 | 0.1 |
| Thigh circ (cm) | 18.1±4.8 | 18.9±4.3 | 0.5 | 18.8±2.7 | 18.7±2.2 | 0.6 |
| Peak VO<sub>2</sub><br>(mL/kg/min) | 22.3±7.1 | 26.1±6.8 | 0.006 | 24.7±8.4 | 26±9.4 | 0.3 |
| Peak VO<sub>2</sub><br>(% predicted) | 60±15.3 | 70.2±17.8 | 0.002 | 69.2±17.3 | 73.7±20.6 | 0.2 |
| 6MW distance<br>(m) | 384.5±118.7 | 488.2±125.0 | <0.0001 | 457.5±89.7 | 496.5±64.8 | 0.04 |
| Sit to Stand (reps) | 15.8±5 | 22.1±6.9 | <0.0001 | 17.8±4.9 | 21.6±6.6 | 0.03 |
| Sit & Reach (in) | 14.7±3.3 | 16.1±3.6 | 0.004 | 14.4±4.2 | 15±3.3 | 0.004 |
| Arm curls (reps) | 17.9±4.4 | 23.0±5.1 | <0.0001 | 19.5±3.7 | 24.8±4.8 | 0.005 |

Data are presented as mean±SD. A paired t-test was performed to evaluate for change between tests. A p-value <0.05 was considered significant.
**Abbreviations:** kg (kilograms), cm (centimeters), m (meters), circ (circumference), VO<sub>2</sub> (oxygen consumption), mL (milliliters), min (minute), VAT (ventilatory anaerobic threshold), VE (ventilatory equivalents for oxygen), VCO<sub>2</sub> (ventilatory equivalents for carbon dioxide), HR (heart rate), bpm (beats per minute), reps (repetitions), in (inches).

When evaluating for changes in mental health and emotional outcomes based on the surveys completed, Duke Activity scores improved significantly in both groups (facility-based 35.0±16.4 v 39.8±13.6, p=0.02; virtual 32.6±16.7 v 36.4±15.8, p=0.04) (Table 5). PCS scores and Rate your Plate significantly improved in the facility-based cohort but not the virtual cohort. There was no significant difference in PHQ-9 and MCS scores for either group.

**Table 5:** Self-reported mental and social outcomes for the patients that completed facility-based (n=33) and virtual (n=12) cardiac rehabilitation.

|  | Facility-based | Facility-based | P value | Virtual | Virtual | P value |
| --- | --- | --- | --- | --- | --- | --- |

|  | Pre-CR | Post-CR |  | Pre-CR | Post-CR |  |
| --- | --- | --- | --- | --- | --- | --- |
| PHQ-9 | 5.6±4.1 | 3.6±3.8 | 0.06 | 6.5±5 | 5.5±4.6 | 0.2 |
| Duke Activity<br>Score | 35.0±16.4 | 39.8±13.6 | 0.02 | 32.6±16.7 | 36.4±15.8 | 0.04 |
| MCS | 49.5±8.4 | 52.2±7.6 | 0.2 | 45.0±13.9 | 46.0±11.8 | 0.9 |
| PCS | 39.5±10.9 | 45.7±8.9 | 0.004 | 41.2±5.9 | 44.8±8.5 | 0.3 |
| Rate your plate | 44.4±8.3 | 47.5±8.4 | 0.006 | 49.1±8.7 | 46.4±7.6 | 0.6 |

Data are presented as mean±SD. A paired t-test was performed to evaluate for change between tests. A p-value <0.05 was considered significant.
**Abbreviations:** PHQ-9 (patient health questionnaire-9), MCS (mental component scoring for vitality, social functioning, emotion health, and mental health), PCS (physical component scoring for physical function, bodily pain, and general physical health).

## Discussion

This study describes the CR outcomes in HD patients during the COVID-19 pandemic utilizing both facility-based and emerging virtual CR options. Despite the difficulties of the COVID-19 pandemic, our pediatric hospital-based cohort demonstrated improvements in functional outcomes evidenced by increases in muscular strength and endurance. Functional strength improved in both the facility-based and virtual CR groups, but peak cardiorespiratory fitness (peak VO_2_) improved more in the facility-based CR group. Additionally, the entire cohort demonstrated improvement in their scales assessing their perceptions of physical and mental health with these changes larger in the facility-based CR cohort.

CR has been shown to be an effective treatment tool to improve muscular strength and endurance in patients with HD^22-23^. As CR expands to more pediatric centers, most congenital cardiac programs have focused on facility-based models of delivering care. The COVID-19 pandemic proved to be a challenge to many CR programs as facility-based care was difficult secondary to COVID-19-based mitigation strategies and disease burden^24^. Additionally, many of the previous exercise intervention studies have been in research populations, that often included participants with a limited variety of cardiovascular diseases, and who were potentially given external financial rewards depending on the study. This study differs in that this CR study was performed on “real world” patients as opposed to research participants.

Muscular endurance (peak VO_2_, 6-minute walk distance) and strength (sit to stand, arm curls) improved in our cohort, which is consistent with other studies involving patients with HD^6, 25-31^. There was an improvement in muscular strength whether care was given facility-based or virtual, which is consistent with previous research^11, 29, 32^. The lack of statistically significant improvement in peak VO_2_ in the virtual CR cohort is noteworthy and likely secondary to several reasons. One possibility for this lack of improvement may be due to the lack of aerobic conditioning equipment. Aerobic fitness equipment such as a cycle ergometer and treadmill are often larger, more expensive, and harder to gain access to compared to strength training equipment. To counteract the lack of access to aerobic equipment, many virtual programs such as our own often rely on high-intensity interval training during the sessions, which attempts to improve muscular endurance through a different mechanism than traditional steady-state aerobic exercise^12,33^. This may point toward high-intensity interval training not being as effective in improving muscular endurance as steady-state aerobic exercise in HD populations. Alternatively, the lack of improvement in peak VO_2_ in the virtual CR group may be due to the overall lower numbers in the virtual cohort or this cohort not having a comparable aerobic intensity and volume with the facility-based cohort. The virtual CR cohort also started with a higher peak VO_2_ and a 6-minute walk distance, so there may also be a ceiling effect in the virtual cohort. Additional studies should be performed in a larger virtual CR cohort to determine if our findings can be reproduced.

Mental health is a critical component of the care of those with HD^34^. Regular exercise has been shown in non-HD populations to improve mental health, dietary habits, sleep hygiene, and quality of life^35-36^. Less is known whether this can be reproduced in children and young adults with HD who engage in CR^37-39^. Kroll et al. were able to demonstrate improved parent perception of social and emotional health but not improved patient social and emotional functioning in a largely pediatric HD cohort^39^. Our cohort of pediatric and young adult HD patients did see an improvement in emotional health as evidenced by a lower PHQ-9 and improved perceived fitness based on improvements in the Duke Activity and Physical Component Scores. Additionally, the facility-based CR cohort noted a significant improvement in their dietary habits, evidenced by the improvement in the Rate your Plate scores. This supports the importance of exercise of the social, cognitive, and emotional health in patients with HD. These improvements were seen more in the facility-based cohort compared to the virtual group. This could be secondary to the lack of facility-based counseling and patient-provider connection in the virtual group. On the other hand, this could be secondary to the smaller numbers in the virtual cohort. This should be confirmed in larger studies.

There was minimal change in body composition in our cohort. Overall, among all participants who completed CR, there was an increase in height and skeletal muscle mass that was more noticeable in the facility-based CR group. This is likely due to expectant pubertal changes as this cohort was younger with more participants under the age of 18 years of age at the onset of CR. Additionally, there was no change in any of the body circumferences measured further pointing to the lack of significant body composition change. The lack of improvement in muscle and fat mass may be secondary to not enough emphasis placed on nutrition during the program with this potentially needing to be further stressed in our program. This is supported by the entire cohort not having any significant change in the Rate your Plate scales. Another possible reason for the lack of change may be related to the underlying body habitus on referral. Some patients with heart disease have sarcopenia and are underweight, which would result in different dietary recommendations and goals than the patient with heart disease and obesity. Future research should evaluate body composition and circumference changes in patients with heart disease stratified by BMI percentile, which we, unfortunately, could not do in our cohort secondary to the low numbers.

### Limitations

The results of this study must be interpreted in light of the study design. This was a single-center retrospective cohort; the findings may not be generalizable to other contexts. Enrollment in CR was voluntary, and thus the individuals studied may be more highly motivated than other HD populations. Additionally, the decision to participate in facility-based versus virtual CR was not random and was impacted by patient preference, geography, other life circumstances, and safety considerations. Future studies with randomization of the groups would reduce this type of error. Another potential limitation of this study stems from the fact that the aerobic components of the facility-based CR and virtual CR were different. There is a possibility that the HIIT exercise performed by the virtual CR cohort was lower in intensity than traditional stationary aerobic exercise completed by those who attended sessions in person. This difference in aerobic activity could also contribute to some differences between the cohorts as well. Lastly, this was not an intent to treat analysis and there is no guarantee that when a patient enrolls in CR that they will complete the program.

## Conclusion

Completion of CR in a pediatric HD center resulted in “real world” improvement in both physical and psychosocial outcomes regardless of whether the program was performed facility-based or virtual during the COVID-19 era. Further research is needed to determine the type and intensity of exercise as well as counseling needed to fulfill the full potential benefits of virtual CR in this population. CR remains a critical yet underutilized component of the care of young patients with HD.

## Data Availability

Data is available upon request to the corresponding author

## Abbreviations

AACVPR: American Association of Cardiovascular and Pulmonary Rehabilitation
CPET: cardiorespiratory exercise test
CR: cardiac rehabilitation
HD: heart disease
MCS: mental component scoring for vitality, social functioning, emotional health, and mental health
PCS: physical component scoring for physical function, bodily pain, and general physical health
Peak VO_2_: peak oxygen consumption
PHQ-9: patient health questionnaire-9

## Acknowledgments

The authors would like to acknowledge The Heart Institute and the Cardiopulmonary Exercise Laboratory at Cincinnati Children’s Hospital for their support in this project.

## Funding

This research received no specific grant from any funding agency, commercial, or not-for-profit sectors. ARO was supported by the Heart Institute Research Core (HIRC) at Cincinnati Children’s Hospital.

## Disclosures

None relevant to the topic of this manuscript.

---

**Figure 1:**
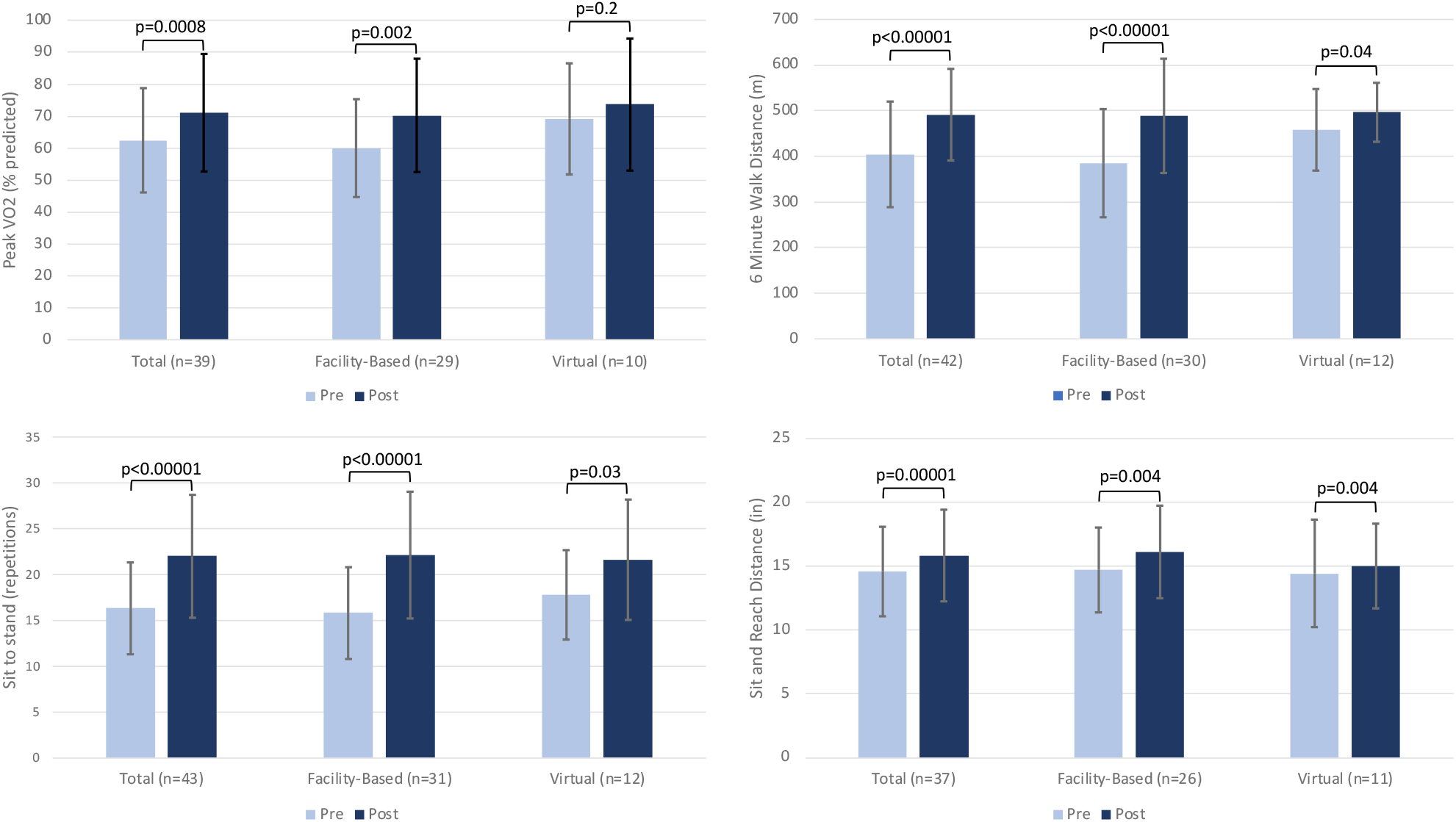
Bar graph demonstrating the changes before and after cardiac rehabilitation in percent predicted peak VO_2_ (1A), 6-minute walk distance (1B), sit-to-stand repetitions (1C), and sit-and-reach distance (1D) for the total cohort, facility-based, and virtual groups. Analysis performed with a paired t-test. **Abbreviations:** VO_2_ (oxygen consumption), m (meters), in (inches)

**Figure 2:**
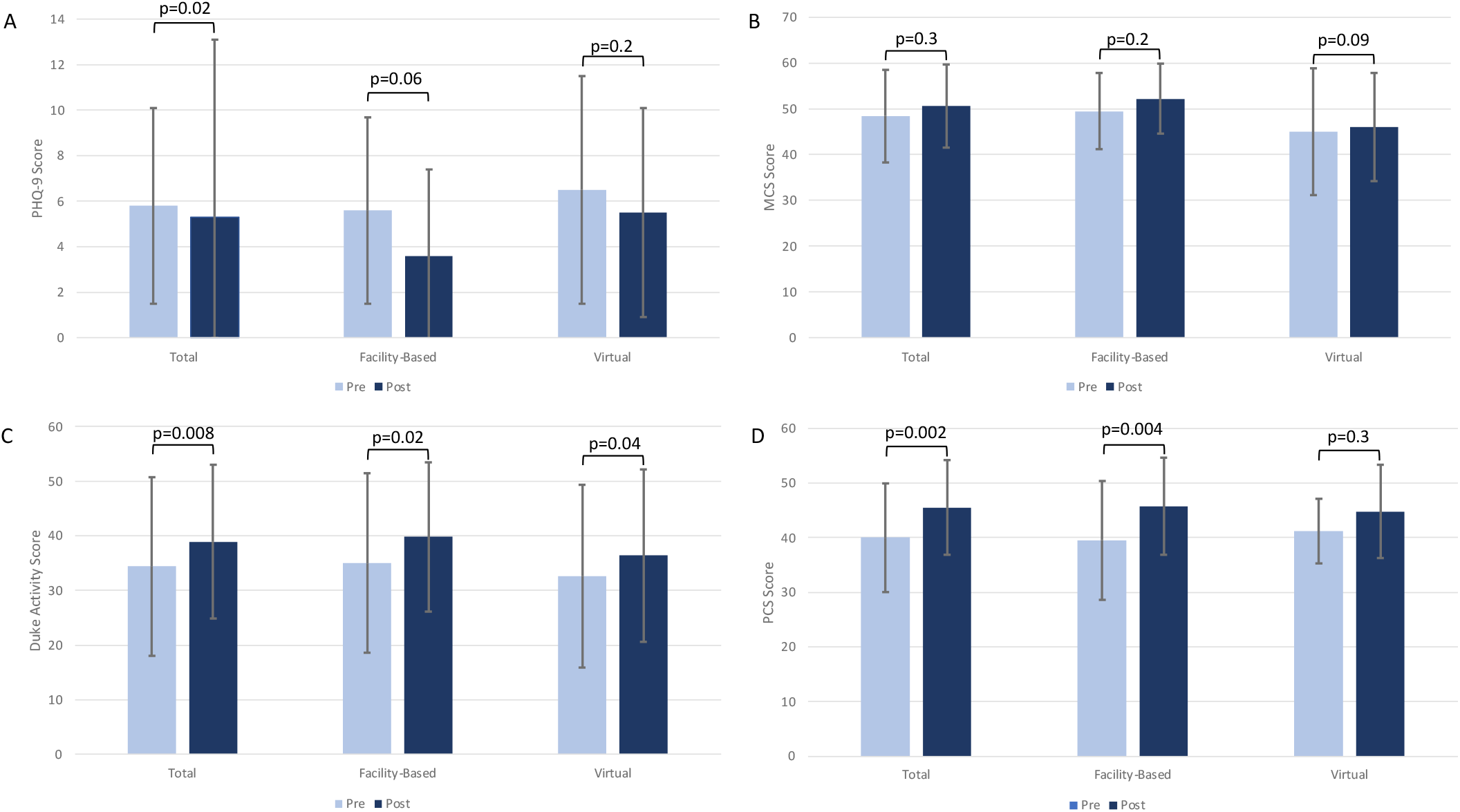
Bar graph demonstrating the changes before and after cardiac rehabilitation in PHQ-9 (2A), mental component (2B), Duke Activity (2C), and physical component scores (2D) in the total cohort, facility-based, and virtual groups. Analysis performed with a paired t-test. **Abbreviations:** PHQ-9 (patient health questionnaire-9), MCS (mental component scoring), PCS (physical component scoring)

## Notes

### Competing Interest Statement

The authors have declared no competing interest.

### Clinical Trial

NA

### Author Declarations

This project was approved by the Cincinnati Children's Hospital Institutional Review Board

